# Association between COVID-19 mortality and population level health and socioeconomic indicators

**DOI:** 10.1101/2021.01.25.21250468

**Authors:** Sasikiran Kandula, Jeffrey Shaman

## Abstract

With the availability of multiple COVID-19 vaccines and the predicted shortages in supply for the near future, it is necessary to allocate vaccines in a manner that minimizes severe outcomes. To date, vaccination strategies in the US have focused on individual characteristics such as age and occupation. In this study, we assess the utility of population-level health and socioeconomic indicators as additional criteria for geographical allocation of vaccines. Using spatial autoregressive models, we demonstrate that 43% of the variability in COVID-19 mortality in US counties can be explained by health/socioeconomic factors, adjusting for case rates. Of the indicators considered, prevalence of chronic kidney disease and proportion of population living in nursing homes were found to have the strongest association. In the context of vaccine rollout globally, our findings indicate that national and subnational estimates of burden of disease could be useful for minimizing COVID-19 mortality.

## Introduction

By the end of 2020, the COVID-19 pandemic has resulted in 81.5 million documented cases and 1.8 million deaths globally (1). The United States has contributed nearly a quarter of these cases and has lost 1 in every 1000 residents to COVID-19 (2). The outbreak has affected all states in the US but with considerable differences in the trajectory and severity of individual outbreaks. Besides this inter- and intra-state geographical variability, the likelihood of adverse outcomes among those infected is reported to be associated with individual’s age, gender, race/ethnicity and underlying health conditions (3-6). An estimated 22% of the global population and 28% of the US have one or more of the underlying conditions that pose increased risk of sever outcomes from COVID-19(7).

Early studies on clinical characteristics of severe outcomes from COVID-19 were reported from China(5, 8), after the first large outbreak in Wuhan, and concurring estimates were subsequently published from UK, France, US and elsewhere (3, 4, 9-12). Guan et al (8) reported that among 1100 of the earliest laboratory confirmed cases of COVID-19 in China, the presence of co-morbidities such as diabetes, hypertension and chronic obstructive pulmonary disease were more prevalent in those with severe outcomes (admission to ICU, requiring mechanical ventilation or death), along with a slightly elevated risk among men and by now well-established risk with increasing age. Using a larger data sample of 45 thousand cases, Deng et al(5) reported that mortality was associated (relative risk (RR) or hazard ratio (HR)) with cardiovascular disease (RR = 6.75, 95%CI = 5.40–8.43), hypertension (HR = 4.48, 95%CI = 3.69–5.45), diabetes (RR = 4.43, 95%CI = 3.49–5.61) and respiratory disease (RR = 3.43, 95%CI = 2.42–4.87, p < 0.001).

A later, more extensive study(9) from the UK linking 17 million cases to 11 thousand deaths also found association between COVID-19 deaths and kidney disease (HR=2.5, 95%CI = 2.3-2.7), diabetes(HR = 1.95, 95%CI = 1.8-2.1), extreme obesity (HR = 1.9, 95%CI = 1.7-2.1) and several other co-morbidities. From a pooled analysis of 75 studies from multiple countries, Popkin et al(12) summarized that individuals with obesity are at increased risk of death (OR = 1.48; 95% CI, 1.22–1.80) hospitalization (OR = 2.13; 95% CI, 1.74–2.60) and ICU admission (OR = 1.74; 95% CI, 1.46–2.08). Based on these findings and the known prevalence of co-morbidities that existed in the population before the emergence of the pandemic, the populations at risk of severe COVID-19 outcomes at county-level in the US(13) and in several countries have been estimated(7). Other studies have examined the associations of socioeconomic characteristics including poverty, income and race/ethnicity (14-16).

Over the past year, public health attempts to reduce transmission largely centered on non-pharmaceutical interventions such as social distancing, face coverings and hand hygiene. In the US, these interventions have had limited success and part of this failure stems from their dependence on collective compliance. The recent availability of high efficacy vaccines gives individuals an additional tool to protect themselves (vaccine supply permitting), and importantly, does not require cooperation from collective public.

The availability of vaccines also implies an opportunity to refocus our efforts at reducing infections from efforts at reducing severe outcomes by prioritizing vaccination for those at a higher risk of severe outcomes. To date such strategies have been largely guided by individual characteristics such as age and occupation. We hypothesize that population level characteristics can also guide the optimal allocation and distribution of vaccines geographically. This points to a potential two-layered approach of first identifying high-risk communities within which high-risk individuals can be prioritized.

Here, we assess the feasibility of the first part of such an approach and evaluate the extent to which the geographical variability of mortality in US can be explained by population characteristics that predate the epidemic. Our outcome of interest is COVID-19 associated mortality rates at county resolutions, which we attempt to model as a function of population health and socioeconomic indicators. An initial set of indicators associated with COVID-19 mortality as reported in peer-reviewed studies, and data sources for estimates of these indicators were identified. A smaller subset of the variables were selected based on the correlation between the variables and their independent effects on the response.

Conventional regression models assume that observations are independent of one another, which in the case of spatial data translates to assuming observations in nearby locations are no more closely related than those farther away. Given the transmission dynamics of COVID-19, counties nearby are likely to be have similar case and death rates and spatial dependence rather than spatial independence is a more appropriate assumption. This spatial dependence also extends to health and socioeconomic indicators and potentially latent and unobservable characteristics that effect mortality.

Spatial autoregressive (SAR) models offer a parsimonious way to augment basic regression models with spatial dependence between locations (17), and are an extensively studied family of analytical approaches with applications ranging from econometrics, environmental studies and health sciences (18-20). In the current study, we first establish the presence of spatial autocorrelation in the response and explanatory variables, thus motivating the need for spatial models. We apply three forms of SAR models, show that they explain a greater proportion of the variability in mortality than linear models and report effect estimates from each.

## Data and Methods

County level indicators of population’s health and social status were retrieved from public sources including the US census and large population surveys. In cases where the survey data are not available at county resolutions, data from prior studies on small-area estimates were used. We tried to limit the number of source dependencies and when alternative estimates were available from multiple sources, we preferred estimates from the US Centers for Disease Control and Prevention (CDC). See Table 1 for a list of sources and descriptions for each variable; Figure 1 presents summary statistics.

**Table 1.**
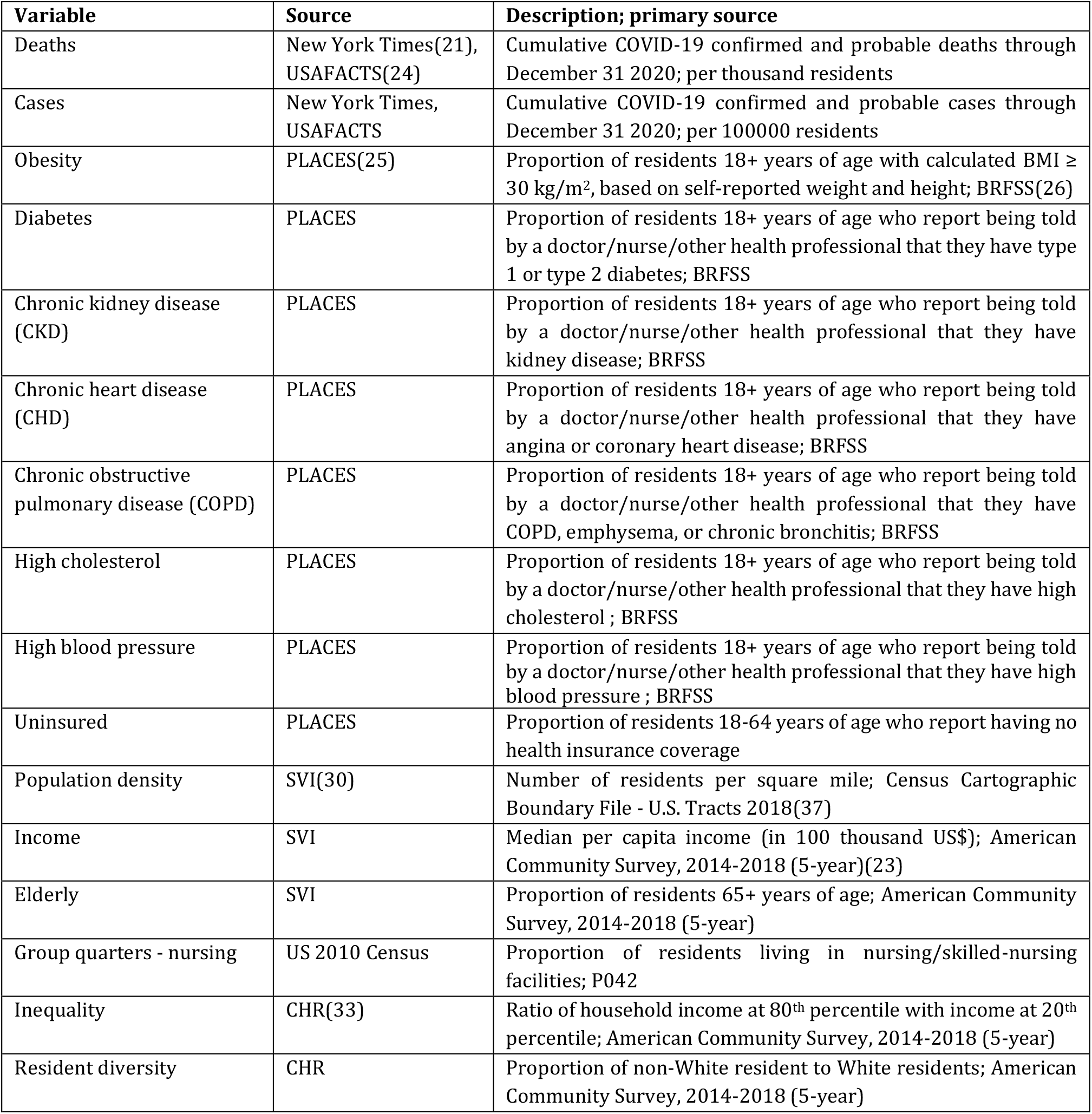
Descriptions and sources for variables included in the study.

| Variable | Source | Description; primary source |
| --- | --- | --- |
| Deaths | New York Times(21), USAFACTS(24) | Cumulative COVID-19 confirmed and probable deaths through December 31 2020; per thousand residents |
| Cases | New York Times, USAFACTS | Cumulative COVID-19 confirmed and probable cases through December 31 2020; per 100000 residents |
| Obesity | PLACES(25) | Proportion of residents 18+ years of age with calculated BMI ≥ 30 kg/m<sup>2</sup>, based on self-reported weight and height; BRFSS(26) |
| Diabetes | PLACES | Proportion of residents 18+ years of age who report being told by a doctor/nurse/other health professional that they have type 1 or type 2 diabetes; BRFSS |
| Chronic kidney disease (CKD) | PLACES | Proportion of residents 18+ years of age who report being told by a doctor/nurse/other health professional that they have kidney disease; BRFSS |
| Chronic heart disease (CHD) | PLACES | Proportion of residents 18+ years of age who report being told by a doctor/nurse/other health professional that they have angina or coronary heart disease; BRFSS |
| Chronic obstructive pulmonary disease (COPD) | PLACES | Proportion of residents 18+ years of age who report being told by a doctor/nurse/other health professional that they have COPD, emphysema, or chronic bronchitis; BRFSS |
| High cholesterol | PLACES | Proportion of residents 18+ years of age who report being told by a doctor/nurse/other health professional that they have high cholesterol ; BRFSS |
| High blood pressure | PLACES | Proportion of residents 18+ years of age who report being told by a doctor/nurse/other health professional that they have high blood pressure ; BRFSS |
| Uninsured | PLACES | Proportion of residents 18-64 years of age who report having no health insurance coverage |
| Population density | SVI(30) | Number of residents per square mile; Census Cartographic Boundary File - U.S. Tracts 2018(37) |
| Income | SVI | Median per capita income (in 100 thousand US\$); American Community Survey, 2014-2018 (5-year)(23) |
| Elderly | SVI | Proportion of residents 65+ years of age; American Community Survey, 2014-2018 (5-year) |
| Group quarters - nursing | US 2010 Census | Proportion of residents living in nursing/skilled-nursing facilities; P042 |
| Inequality | CHR(33) | Ratio of household income at 80<sup>th</sup> percentile with income at 20<sup>th</sup> percentile; American Community Survey, 2014-2018 (5-year) |
| Resident diversity | CHR | Proportion of non-White resident to White residents; American Community Survey, 2014-2018 (5-year) |

**Figure 1.**
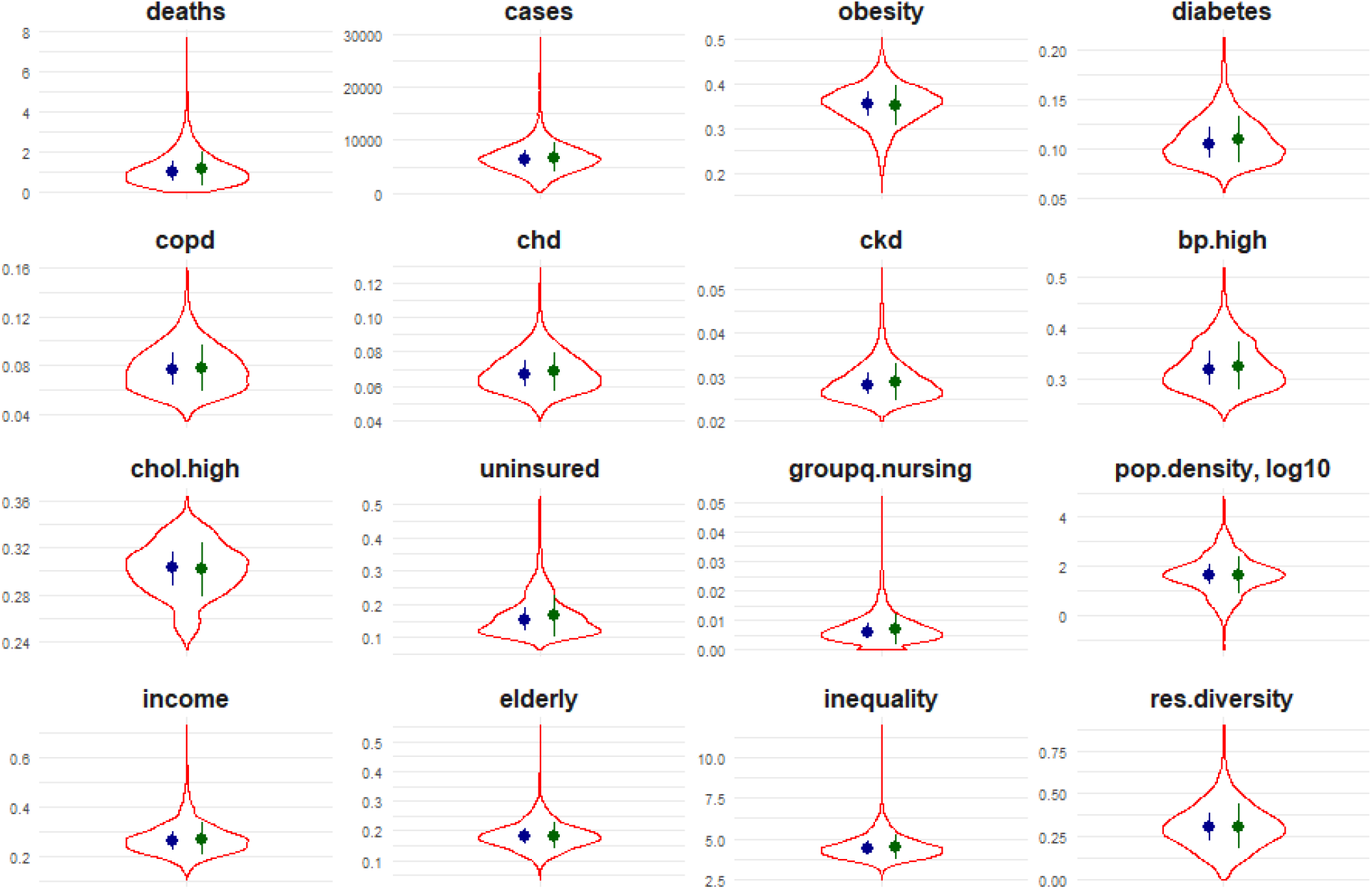
Violin plots of distribution for each variable among counties in the US, along with median (interquartile range) in blue and mean (standard deviation) in green.

### New York Times

Counts for cumulative cases and deaths through December 31, 2020 were retrieved from New York Times public repository (21). These data included both confirmed and probable cases and deaths at the US county-level and is based on Times’ monitoring and analyses of news conferences, data releases and communications with public officials. The determination of cases and deaths as either confirmed or probable is made per definitions laid out in the position statement of the Council of State and Territorial Epidemiologists (22). But as the application can vary across local agencies, here we treat both confirmed and probably categories identically and use total cases and deaths. Case and death rates as a proportion of residents are based on county population estimates from the American Community Survey (ACS) 2014-2018(23).

County-specific data for the 5 counties in New York City were retrieved from USAFACTS (24) as the Times’ data source was found to combine counts for these five counties into a single entity. Figure 2 shows maps of reported county case and death rates.

**Figure 2.**
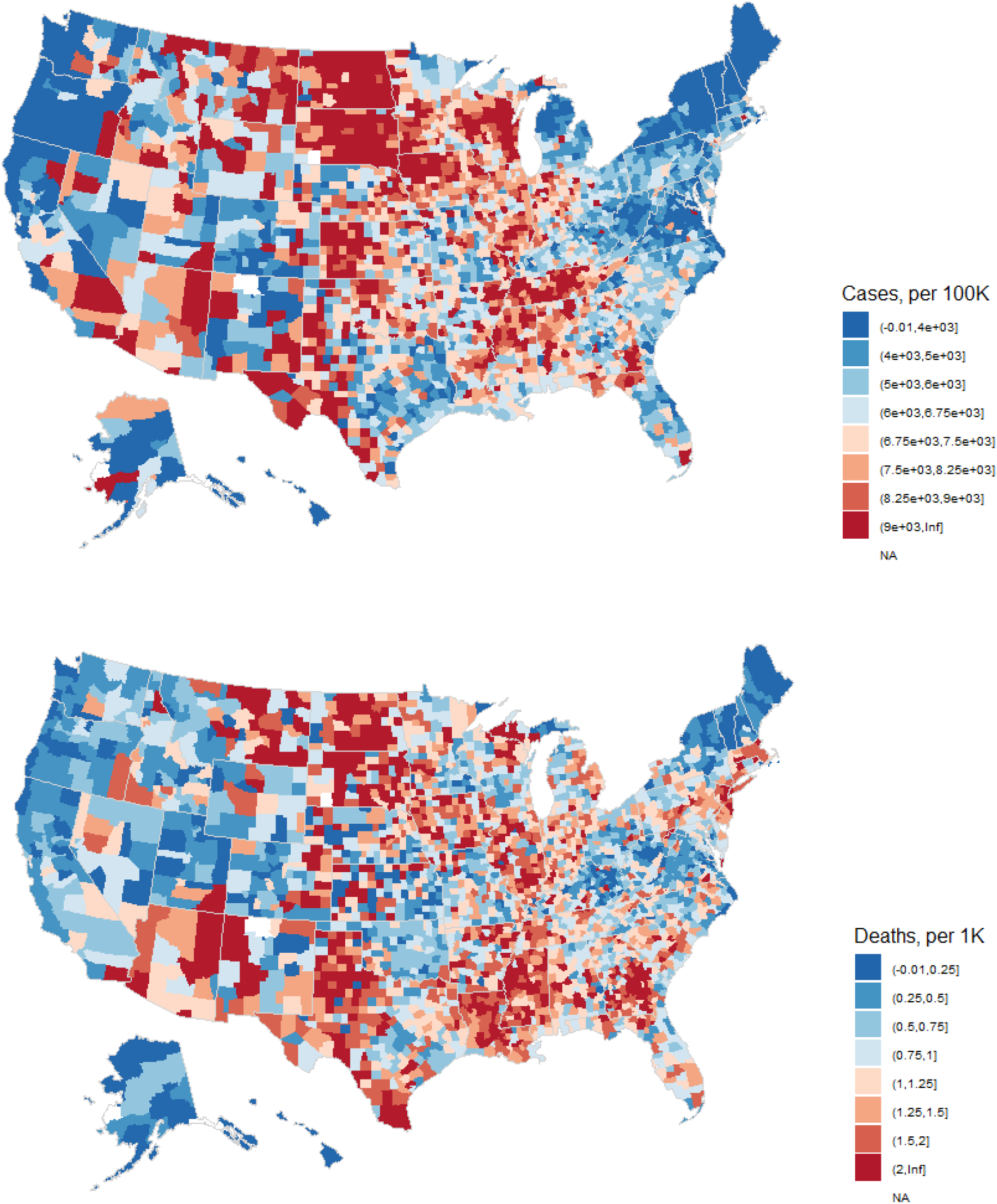
COVID-19 cases (per 100000 residents) and deaths (per 1000 residents) in US counties through December 31, 2020.

### Population Level Analysis and Community Estimates (PLACES)

From the PLACES study(25), a collaboration between the CDC and Robert Wood Johnson Foundation, estimates for population-level health and behavioral indicators were retrieved. These small-area estimates of population health outcomes across the US at county resolutions were generated using data collected through the Behavioral Risk Factor Surveillance System (BRFSS)(26), the US decennial 2010 census and the ACS, following a multi-level regression and post-stratification approach (27, 28).

Of the 27 indicators available in PLACES, we extracted 5 measures of population level prevalence of health conditions that are reported to have individual level associations with COVID-19 outcomes, namely obesity, diabetes, chronic obstructive pulmonary diseases and chronic heart and kidney diseases. In addition, three related health indicators, the prevalence of high blood pressure and high cholesterol and proportion of residents uninsured were also included.

### Social Vulnerability Index

CDC’s Social Vulnerability Index (SVI) is a measure of a county’s relative vulnerability to hazardous events (29, 30) and is intended to help public officials and planners better prepare for such events. Overall, county ranks are based on fifteen socioeconomic indicators collected in the ACS. Three of the factors in the SVI, namely county population density, median per capita income and proportion of the population that is older than 65 years of age, are hypothesized to be associated with COVID-19 mortality (14, 15, 31, 32). As association between the other variables in SVI and COVID-19 is uncertain, we limited inclusion to the raw estimates of these 3 variables and ignore the other variables in SVI and the overall index.

### County Health Rankings

Two additional variables derived from the ACS 2014-18 and available through the County Health Rankings (CHR)(33), are hypothesized to be measures of socioeconomic disparities in a county and included in this study: ratio of the 80^th^ percentile income to 20^th^ percentile as a measure of income inequality, and the proportion of non-White to White residents as a measure of racial diversity. We observed that estimates for these variables in a small percentage (∼1.5%) of counties were missing and used the following three-step process to impute missing values: a) the mean of neighboring (defined in later sections) counties that have estimates; b) if there are no neighbors with estimates, the median of all counties in the state for which estimates are available; c) if estimates are missing for all counties in a state, the median across all counties in the US for which estimates are available.

### US 2010 Census

It has also been reported that COVID-19 clusters occur in facilities in which people live in group quarters, where the increased vulnerability can result from either the living conditions in such facilities (difficulty to social distance in correctional facilities or on college campuses, for example) or the characteristics of the residents (elderly in nursing homes with underlying health conditions)(34, 35). As mortality from COVID-19 is known to be less likely in younger populations, we focused instead on elderly living in group quarters. An estimate of proportion of the population living in nursing homes or facilities with skilled-nursing in each county was included in this analysis (Table 1).

#### Methods overview

We first built linear univariate models for each predictor with county-level COVID-19 mortality as outcome, adjusting for county case rates. These models inform both the individual effects and the proportion of variance in mortality explained by each of these predictors. We followed this with a linear multivariate model, again adjusting for case rates. In both univariate and multivariate models, observational independence is inappropriate because of spatial autocorrelation in both the response and predictors. We verify this by standard tests on the residual of the multivariate model. We finally build spatial simultaneous autoregressive models and report effect estimates.

### Spatial weight matrix and spatial autocorrelation

As introduced in an earlier section, a key assumption in standard ordinary least square regression (OLS) models is the independence of observations that does not hold because COVID-19 cases and deaths in a county are related to cases and deaths in other counties (spatial dependence) and often counties adjacent to it (spatial autocorrelation). Models that do not account for spatial dependence and autocorrelation are shown to have inflated type I errors (20, 36).

To establish adjacency of counties in the US, we define a simple spatial *n x n* matrix, **W**, using shape files that list county boundaries as an ordered set of geocoded reference points (37). County adjacency is defined by queen congruity (at least one shared boundary point) and the spatial weight matrix is row standardized i.e. for each county *i*, the weight of link to county *j, w*_*ij*,_ is the inverse of the number of neighbors of *i*, if *j* is adjacent to *i*, and 0 otherwise; ∑_*j*_ *w*_*ij*_ =1. A county is assumed to not be a neighbor of itself i.e. *w*_*ij*_ *= 0* when *i =j*.

Moran’s I(38, 39), a commonly used measure of global spatial autocorrelation, is calculated as:

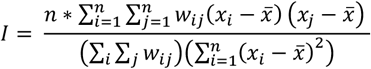

where *n* is the number of counties, *x*_*i*_ is the variable of interest for county *i*, 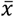 is the mean across all counties and *w*_*ij*_ is as defined by the spatial weight matrix, **W**. Here, as ***W*** is row standardized ∑_*i*_ ∑_*j*_ *w*_*ij*_ = *n* and the above equation can be simplified. The significance of the statistic was tested under the randomization assumption i.e. *x*_*i*_ are draws from a random distribution and there is no spatial association. A related measure to identify specific regions within the study region that exhibit spatial autocorrelation, the Local Moran’s I, was also estimated. Figure 3 shows Moran’s I and counties with significant(40) Local Moran’s I and for each predictor and outcome.

**Figure 3.**
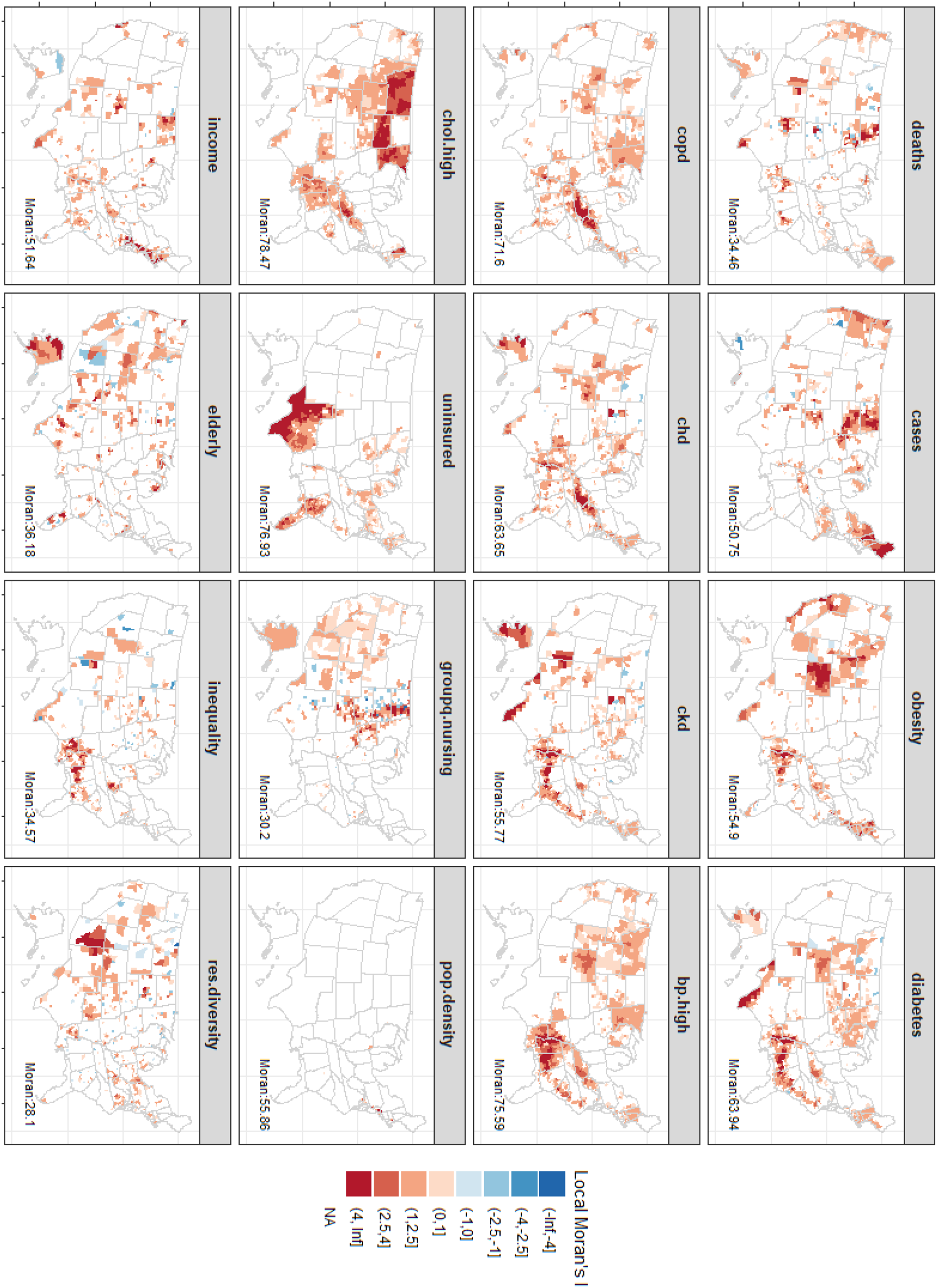
Local Moran’s I statistic for spatial autocorrelation for all measures and outcome. Only counties where the statistic is significant (p < .05) are shown. Significance is tested under Pr[I – E(I)/Var(I)] as given by Anselin(40). Global Moran’s I statistic is denoted by the label in each subpanel and found to be statistically significant for all variables.

We were also interested in determining whether spatial autocorrelation, if present, resided in the response or in the residual, as this also informs the choice of the spatial model. To identify this we used robust Lagrange Multiplier tests that can detect possible autocorrelated residuals in the presence of an omitted lagged response and vice versa (41, 42). The statistics reported here are from implementations of these tests in the *spdep* (39, 43) R(44) library.

### Variable pruning

As the variables selected for inclusion are related, we calculated Spearman’s correlation between pairs of variables (Figure 4) and found some of the variables to be very highly correlated. Hence, it would not be appropriate to include these pairs together in models. We used the results of the univariate analysis, to aid variable selection by only retaining those variables that have a correlation of less than .75 with variables of a higher R^2^. This led to the elimination of five variables – prevalence indicators for diabetes, heart disease, high blood pressure (all highly collinear with kidney disease), high cholesterol (collinear with COPD) and median per capita income. The linear multivariate model and the spatial models were built using this smaller set of predictors (n = 9).

**Figure 4.**
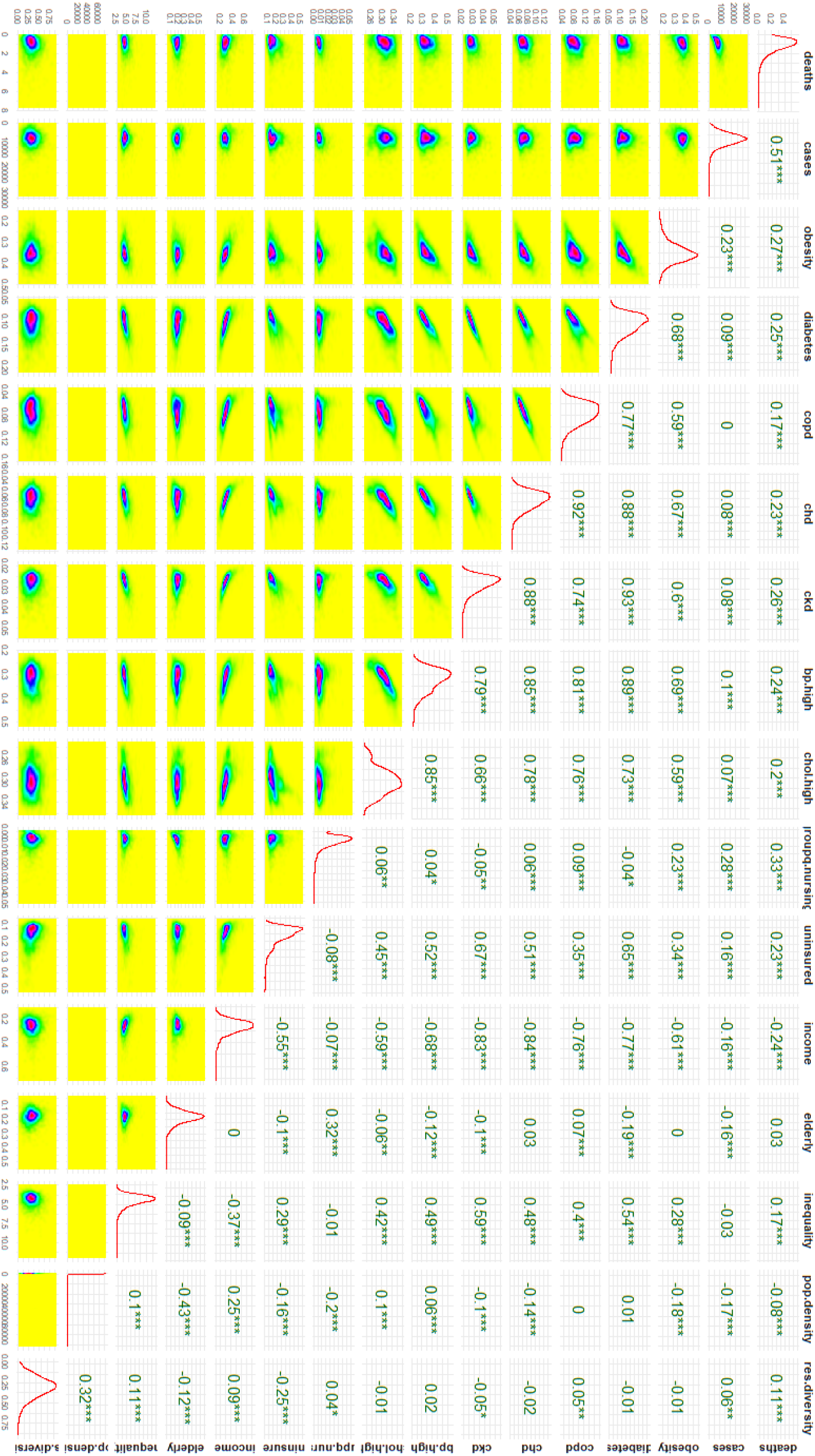
Pairwise surface plots (below diagonal), Spearman correlation (above diagonal) and density (diagonal) of outcome and measures used in the study. * indicates level of statistical significance of the correlation: *p*<0.001 (***); 0.001≤ *p* < 0.01 (**); 0.01≤ *p* < 0.05 (*).

### Spatial simultaneous autoregressive models (SAR)

The general form of an autoregressive model in spatial statistics is given by (17, 20, 45):

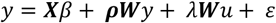

where *y* is a *n ⨯ 1* vector of the response variable, **X** is a *n ⨯ k* matrix of *k* predictors for *n* counties, **W** is the *n ⨯ n* spatial weight matrix, *ρ* is the spatial autoregressive *lag* coefficient and λ the spatial *error* coefficient and *β, u* the coefficient and error vectors respectively. When λ = 0, the autoregressive process is assumed to occur in the response only (captured by ρ**W**) and the model is referred to as a spatial lag model. When ρ = 0, the autoregressive process is assumed to occur only in the errors (captured by λ**W**), and the model referred to as spatial error model. Model implementations are per *spatialreg (43, 45, 46)* library in R.

## Results

Results from the univariate analysis indicate that the selected variables individually explain 24-29% of the variability in mortality, adjusting for case rates. Mortality is estimated to increase by 43 per thousand residents (95% CI: 37-49) for every 1% increase in prevalence of chronic kidney disease, and by 10.4 (95% CI: 8-13) for chronic heart disease, 7.4 (95% CI: 6-8) for diabetes, 4.4 (95% CI: 3-5.8) for COPD, 3.7 (95% CI: 2.6-5.8) for high cholesterol, 2.8 (95% CI:2.2-3.3) for high blood pressure and 2.6 (95% CI: 2-3.2) for obesity prevalence respectively (Figure 5). These health indicators also explain 28%, 25.5%, 27.5%, 24.6%, 24.6%, 25.9% and 25.3% of the variability respectively.

**Figure 5.**
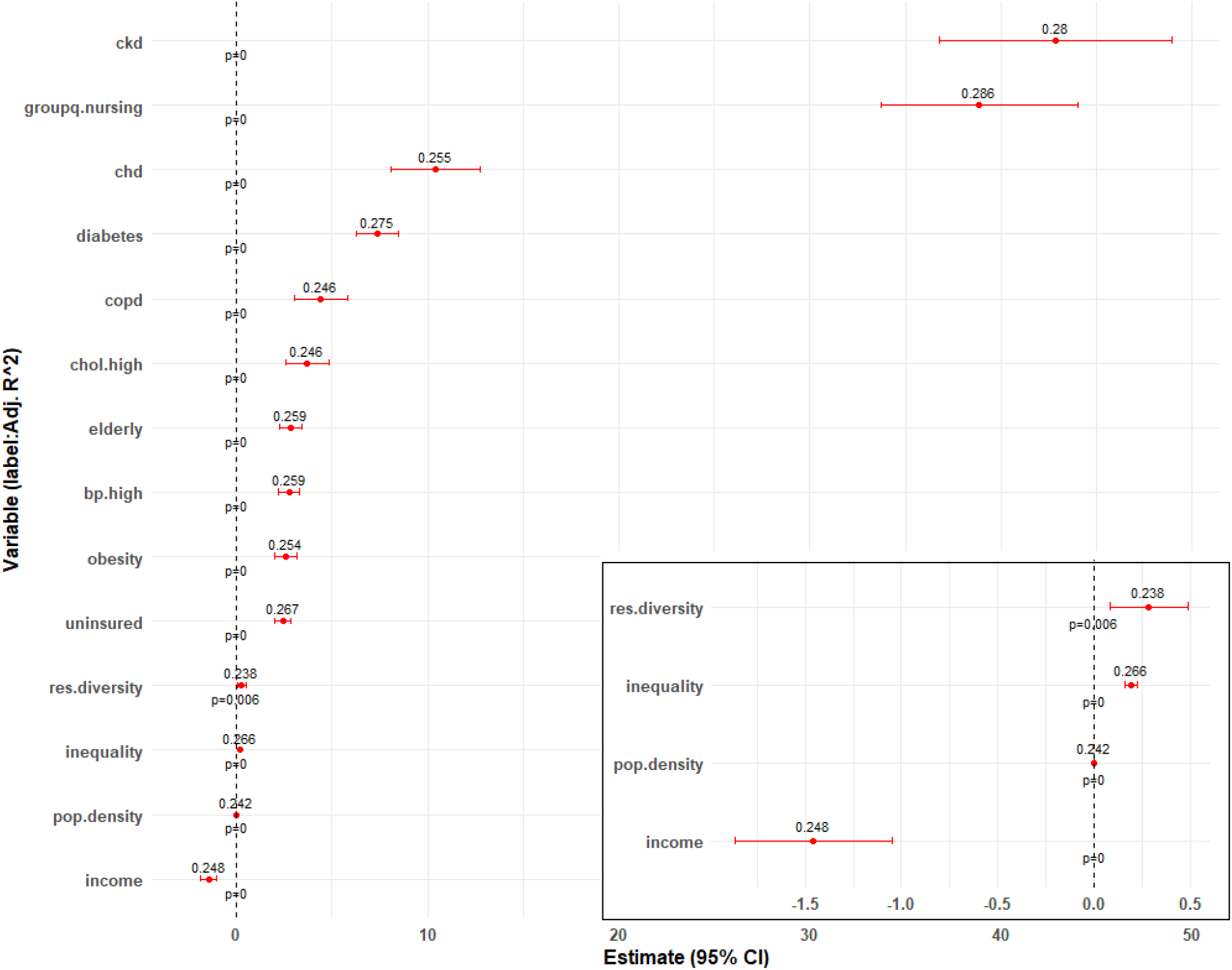
Estimates (95% CI) of health and socioeconomic indicators in a linear univariate model with death rate as outcome and adjusting for COVID-19 case rates. Labels indicate adjusted R^2^. Inset magnifies select variables of smaller estimates.

Among socioeconomic indicators, the largest association was seen with the nursing home variable (Adjusted R^2^: 29%) with an estimated increase of 39 deaths per thousand (95% CI: 34-44) for every 1% increase in percent living in nursing homes. Mortality rates are estimated to increase by 2.8 (95%CI: 2.3-3.4) and 2.4 (95% CI: 2-2.9) for each 1% increase in percentage of the population who are elderly (65+ years) and uninsured 18-64 year olds, respectively. In contrast, mortality rate is estimated to decrease by 1.5 (95% CI: 1.05-1.87) for every thousand dollar increase in per capita income. On average, the R^2^ estimates for socioeconomic indicators are lower than for health indicators.

Following variable pruning to correct for collinearity, the multivariate model explained 38% of the variability in mortality with a few changes in effect estimates. Obesity’s association is not statistically significant in the presence of kidney disease and COPD’s association is counterintuitively negative (Table 2).

**Table 2.**
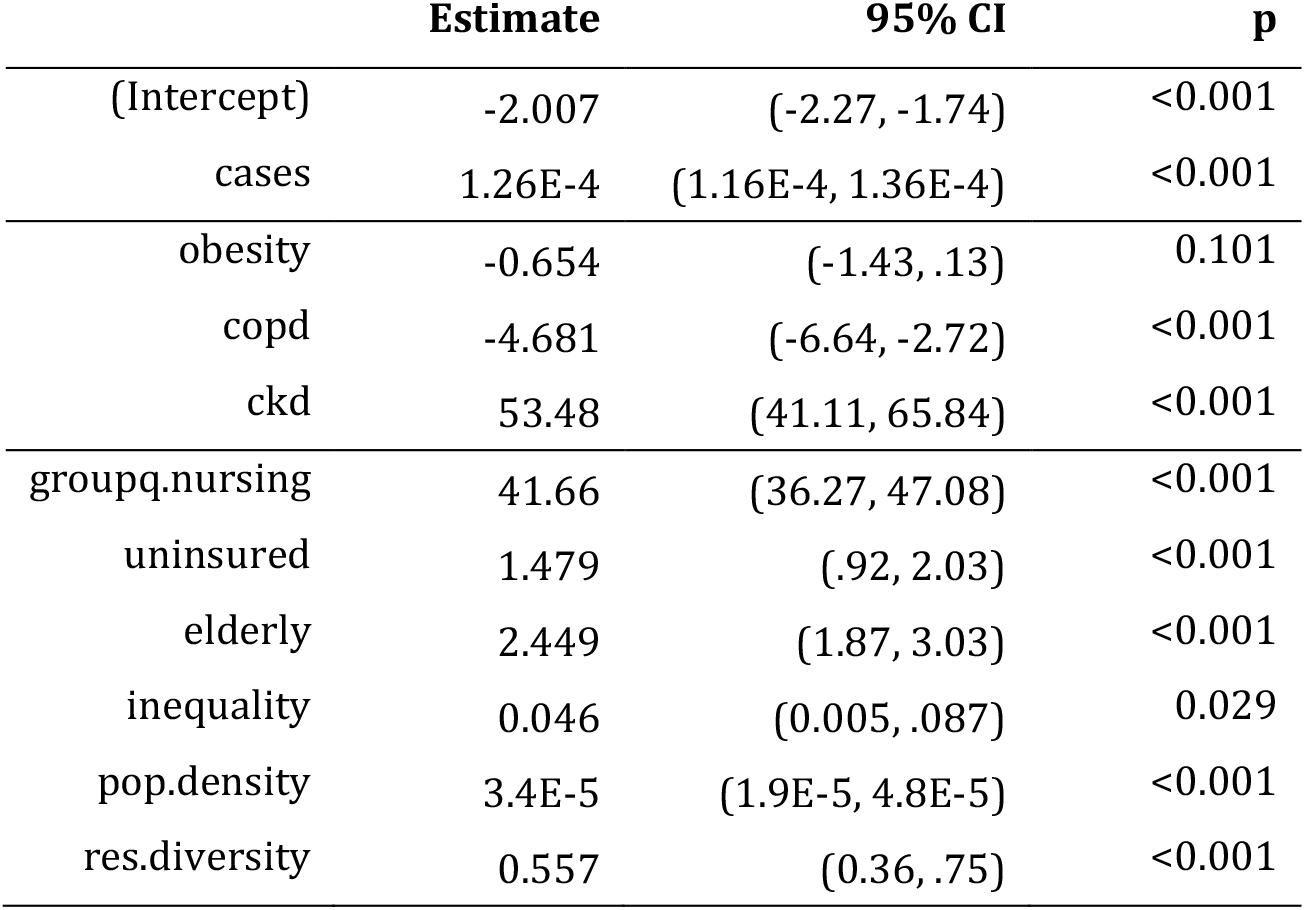
Results of multivariate analysis with linear model, adjusting for case rate. Adjusted R^2^ = .3812; F-statistic = 194. (p-value: < .001)

|  | Estimate | 95% CI | p |
| --- | --- | --- | --- |
| (Intercept) | -2.007 | (-2.27, -1.74) | <0.001 |
| cases | 1.26E-4 | (1.16E-4, 1.36E-4) | <0.001 |
| obesity | -0.654 | (-1.43, .13) | 0.101 |
| copd | -4.681 | (-6.64, -2.72) | <0.001 |
| ckd | 53.48 | (41.11, 65.84) | <0.001 |
| groupq.nursing | 41.66 | (36.27, 47.08) | <0.001 |
| uninsured | 1.479 | (.92, 2.03) | <0.001 |
| elderly | 2.449 | (1.87, 3.03) | <0.001 |
| inequality | 0.046 | (0.005, .087) | 0.029 |
| pop.density | 3.4E-5 | (1.9E-5, 4.8E-5) | <0.001 |
| res.diversity | 0.557 | (0.36, .75) | <0.001 |

Moran’s I test for spatial autocorrelation in residuals of the above model was found to be statistically significant (18.4, *p* < 1E-6). Both robust LM tests were found to be significant indicating possible autocorrelation in both the error (28.7, *p* < 1E-6) and response (33.5, *p* < 1E-6). Hence, three model forms, the general SAR model, spatial lag and spatial error models were attempted.

The proportion of variability explained by the SAR models is about 14% higher than the linear model (Figure 6). The spatial error model had an Nagelkerke R^2^ (47) of 43.5% with an estimated autocorrelation error coefficient (λ) of .418 (95%CI: .37 - .46). The spatial lag model and the general model were observed to have an R^2^ nearly identical to that of the error model, The autocorrelation coefficient in response (ρ) was found to be .347 (95%CI: .31-.39) for the spatial model, but when both coefficients were estimated simultaneously in a general model, the lag coefficient was found to be not significant: λ = .336 (95%CI: .244-.429); ρ = .083 (95%CI:-.007 - .174; *p*=.07). Figure 7 shows the spatial lag model’s estimates and residuals.

**Figure 6.**
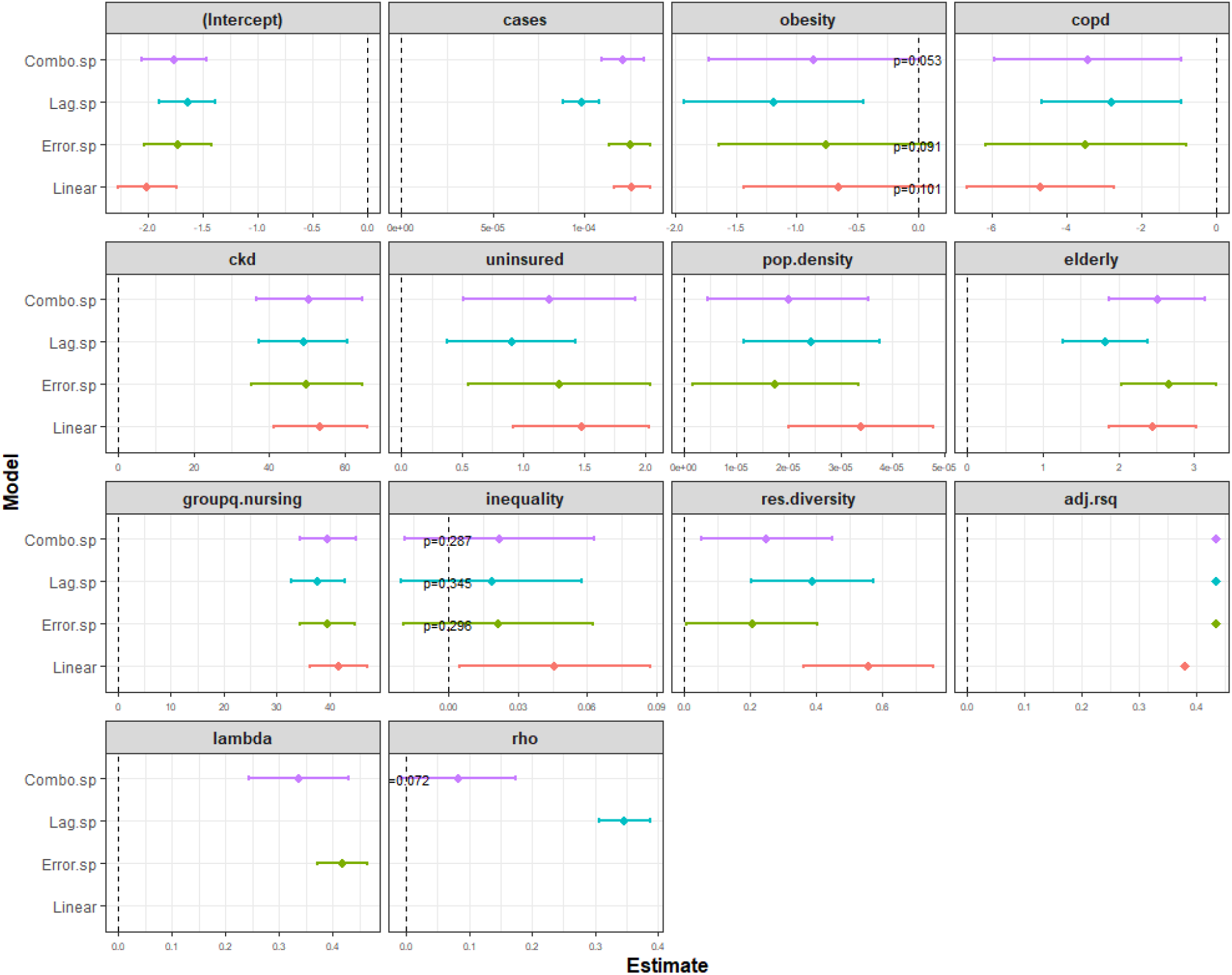
Variables estimates with linear and three spatial regression models. *p*-values indicated when *p* > .05.

**Figure 7.**
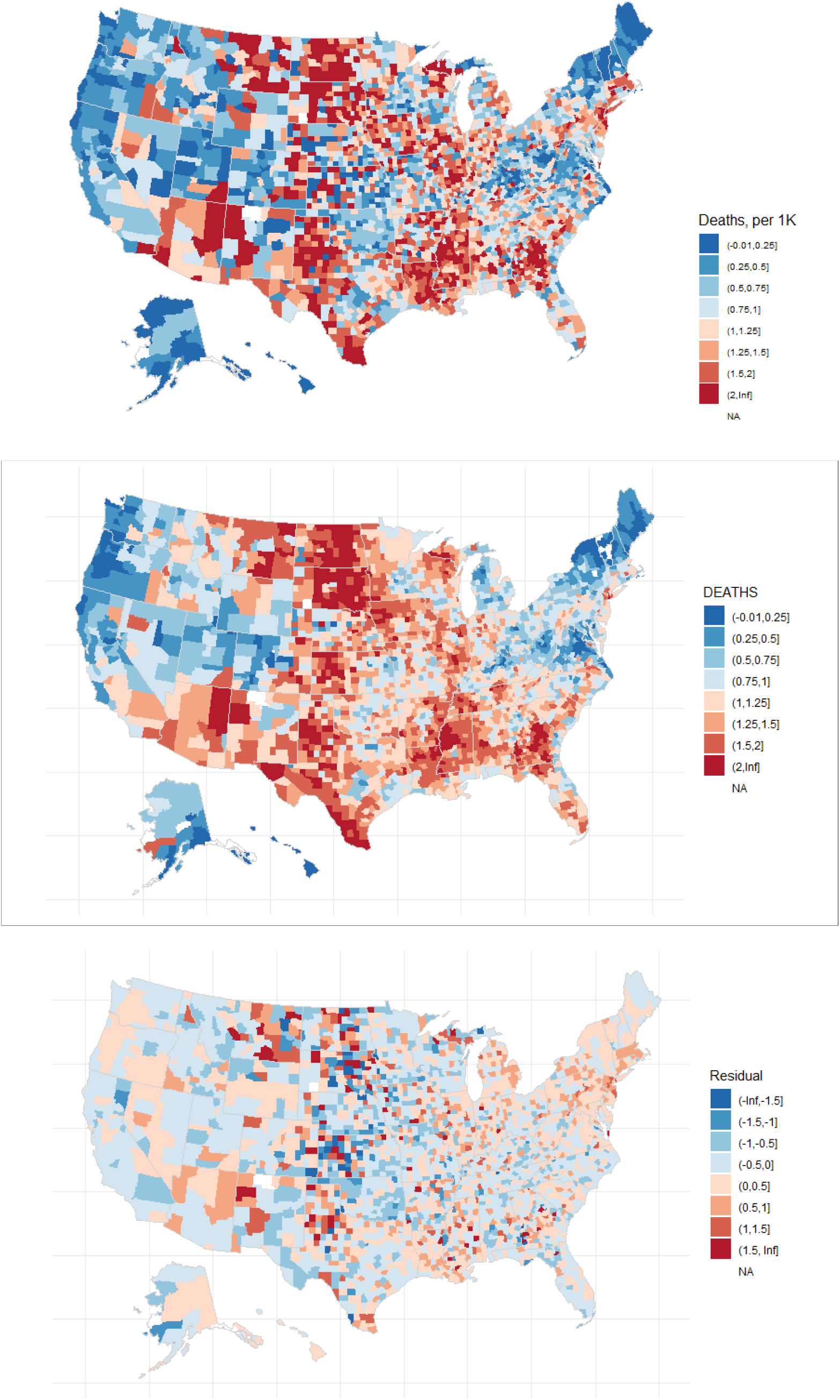
Observed death rate (as in Figure 2), model fit and residual of the spatial lag model.

The Global Moran’s test on the residuals of all three models found no significant spatial autocorrelation (*p* > .05). Effect estimates for inequality variable were found to be not significant (*p* > .05) in any of the spatial models (Figure 6). The negative association of COPD seen in the linear model is also observed with the spatial models. Obesity, as in the linear model, was found to be not statistically significant in two of the spatial models.

To test for sensitivity of models’ R^2^ to the variable pruning method, we additionally subset variables using alternative spearman correlation thresholds of .5, .65 and .85 and built linear and spatial models with each. Figure 8 shows that R^2^ was not sensitive to the value of threshold and the spatial models have a consistently higher R^2^ than the linear model.

**Figure 8.**
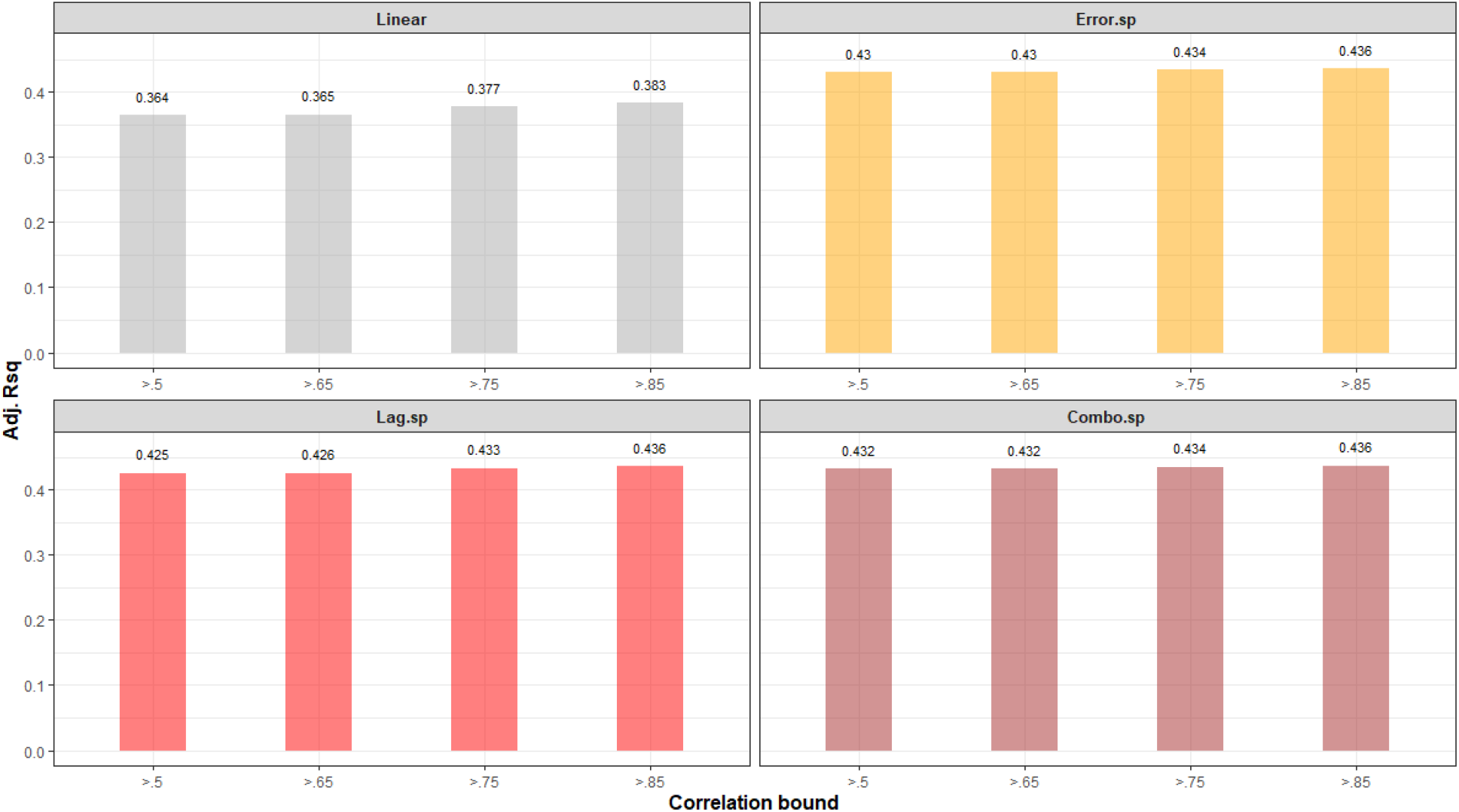
Sensitivity of Adhusted R^2^ to Spearman correlation threshold used in variable pruning.

## Discussion

We have built models to estimate COVID-19 mortality rates for given case rates and population health and socioeconomic characteristics. Our results indicate that together these indicators can explain 43% of the variability in US county mortality rates, when spatial autocorrelation is accounted for. We found that among health indicators considered the prevalence of chronic kidney disease and among socioeconomic indicators the proportion living in nursing homes, have the largest associations with mortality.

The choice and timeliness of control strategies in response to an outbreak do affect its progress and caseload. Our findings here show that differential risks of severe outcomes from COVID-19 across populations can be in part estimated from the structures and contexts in which the outbreak occurs, for example, a population’s quality of health, its access to healthcare and the disparities therein. With the availability of vaccines, these population level indicators can serve as criteria for prioritizing geographical allocation of vaccines.

These findings may also be relevant to low- and middle-income countries (LMIC). It has been reported that almost all of the Pfizer-BioNTech and Moderna vaccine doses to be manufactured through the end of 2021 have been purchased and are reserved for distribution in the US, Canada, UK and the EU (48, 49). Of the 42 countries that have rolled out vaccines by early January 2021, only 6 are middle-income countries and none are low-income countries (50). The COVAX initiative with participation from governments of several LMIC countries, the WHO and partner non-governmental organizations, aims to achieve equitable and affordable access to vaccines globally through a common vaccine purchase and allocation framework (51). When allocation decisions need to span multiple countries, national and subnational socioeconomic indicators and burden of disease estimates can potentially be leveraged to reduce overall risk of severe outcomes from COVID-19 as our findings demonstrate.

This study has a few limitations. Case and death counts were retrieved a week after the end of the study period. Given the lags in data reporting, particularly with deaths, events occurring at the end of the study period may not have been recorded and the rates used are underestimates. Similarly, the outcomes may not yet be known for cases recorded near the end of the study period.

The adjacency based spatial weight matrix that was used in this study does not sufficiently capture the spread of COVID-19. Cases that occur in a county are not only correlated with those in counties geographically adjacent to it but also with counties with which it has strong population mixing; for example, counties with metropolitan centers into which commuters travel from the suburbs, or counties with major airports. Spatial weight matrices that capture mobility patterns may be more appropriate and lead to better spatial models. Similarly, methods that can explicitly account for spatial autocorrelation in predictors remain to be explored.

Finally, the model structure presented may not be parsimonious in the number of predictors. Although we dropped a third of the predictors initially considered (to correct observed collinearity), model forms with a smaller subset of independent variables may yield near identical R^2^ and need to be explored. This is also belied by the lack of significance of some of the predictors included in the spatial models. One approach could start with a minimal set of predictors, incrementally add predictors while evaluating goodness of the resulting model in each iteration and terminating when the improvement is below a threshold. Similarly, the variable pruning discussed above is ad hoc; the variables included in the model may be interchangeable with those discarded with only marginal change in model performance.

## Data Availability

All data are publicly available and referenced in the manuscript.

